# Making maternity and neonatal care personalised in the COVID-19 pandemic: results from the Babies Born Better Survey in the UK and the Netherlands

**DOI:** 10.1101/2022.04.11.22273702

**Authors:** Lauri M.M. van den Berg, Naseerah Akooji, Gill Thomson, Ank de Jonge, Marie-Clare Balaam, Anastasia Topalidou, Soo Downe, the ASPIRE COVID-19 research team (details below)

**Affiliations:** Amsterdam University Medical Centre, Vrije Universiteit Amsterdam, Department of Midwifery Science, AVAG/Amsterdam Public Health, Amsterdam, the Netherlands; Lancashire Clinical Trials Units, University of Central Lancashire, Preston, United Kingdom; School of Community Health & Midwifery, Faculty of Health and Care, University of Central Lancashire, Preston, United Kingdom; Amsterdam University Medical Centre, Vrije Universiteit Amsterdam, Department of Midwifery Science, AVAG/Amsterdam Reproduction and Development, Amsterdam, the Netherlands; University of Central Lancashire; Newcastle upon Tyne Hospitals NHS Foundation Trust; University of Manchester; Amsterdam University Medical Center; University of Southampton; NHS England and NHS Improvement North West; Royal Free Teaching Hospital in London; Birthrights; Kings College London; The University of Nottingham; East Lancashire Hospitals NHS Trust; FivexMore; MVP BAME Group; The Miscarriage Association; RCM; International Confederation of Midwives (ICM); Expert; postnatal care; Royal College of Obstetricians and Gynaecologists (RCOG); Fathers included/Family included/the Family Initiative; Neighbourhood Midwives; Erasmus MC; University of Liverpool; National Maternity Voices; SANDS; Burnet Institute; Australia; National Perinatal Epidemiology Unit (NPEU); London Ambulance Service Trust; University College London; NHS England; Grace in Action; Twins Trust; UCL & City University; Midwifery Unit Network; RCM Scotland; BirthWise NI; British Maternal and Fetal Medicine Society; Birth Trauma Association

## Abstract

**Background:** The COVID-19 pandemic had a severe impact on women’s birth experiences. To date, there are no studies that use both quantitative and qualitative data to compare women’s birth experiences before and during the pandemic, across more than one country.

**Aim:** To examine women’s birth experiences during the COVID-19 pandemic and to compare the experiences of women who gave birth in the United Kingdom (UK) or the Netherlands (NL) either before or during the pandemic.

**Method:** This study is based on analyses of quantitative and qualitative data from the online Babies Born Better survey. Responses recorded by women giving birth in the UK and the NL between June and December 2020 have been used, encompassing women who gave birth between 2017 and 2020. Quantitative data were analysed descriptively, and chi-squared tests were performed to compare women who gave birth pre- versus during pandemic and separately by country. Qualitative data was analysed by inductive thematic analysis.

**Findings:** Respondents in both the UK and the NL who gave birth during the pandemic were as likely, or, if they had a self-reported above average standard of life, more likely to rate their labour and birth experience positively when compared to women who gave birth pre-pandemic. This was despite the fact that those labouring in the pandemic reported less support and choice. Two potential explanatory themes emerged from the qualitative data: respondents had lower expectations during the pandemic, and they appreciated that care providers tried hard to personalise care.

**Conclusion:** Our study implies that many women labouring during the COVID-19 pandemic experienced restrictions, but their experience was mitigated by staff actions. However, personalised care should not be maintained by the good will of care providers, but should be a priority in maternity care policy to benefit all service users equitably.

## Introduction

Pregnancy is a unique and life-changing event that is associated with new experiences and strong emotions for many women and birthing people (further ‘women’) [1, 2]. Events during pregnancy, childbirth and the post-partum period can continue to affect women for many years to come [3]. The onset of the novel coronavirus disease (COVID-19) pandemic has had a severe impact on maternity care provision around the world. More generally, the unknown situation, fear of COVID-19 and a range of public health measures, such as lockdowns and several restrictions influenced the psychological wellbeing of whole populations [4-7]. Besides these stressors for the general population, women in the perinatal period faced additional consequences, such as restrictions in visiting their new-born baby, their place of birth choice, and options for pain medication, that has negatively influenced the mental health of many [8-10].

Over the past two years several studies have evaluated the birth experiences of women during the COVID-19 pandemic. For instance, an Italian qualitative study concluded that pregnant women during the pandemic experienced fear, sadness, and uncertainty when they visualised their upcoming birth [11]. A UK qualitative study of women labouring and giving birth during the pandemic revealed that participants were concerned about feeling alone during birth due to companionship restrictions; the possibility of transmission of COVID-19 to their baby; and restrictions on the use of water for labour and/or birth [12]. In contrast, another Italian study showed similar satisfaction with birth experience for those labouring and giving birth before and during the pandemic [13]. The survey did not explore the reasons for the responses given at both time points. There is therefore a gap in understanding what might influence the experience of maternity care both before and during a system-wide crisis like a pandemic.

To address this gap and provide an in-depth understanding of women’s birth experiences during the COVID-19 pandemic, we used qualitative and quantitative data to compare the experiences of women and childbearing people who gave birth in the UK or the NL either before or during the pandemic. The UK and the NL are European countries with comparable maternity care systems, and, by population size, experienced similar numbers of COVID-19 infections. Their approach to the pandemic in maternity care was somewhat different, with the NL having a more community-based approach and the UK offering more hospital-centric care, but having greater structures in place for service user organisations input [14]. A comparison between the two countries may give insight into how different approaches to the pandemic are related to women’s birth experiences.

This study was undertaken as part of the ESRC/UKRI funded ASPIRE-COVID-19 study, which included an analysis of responses to the international Babies Born Better survey version 3 (B3-survey). The survey includes both quantitative questions about labour and birth experiences, and qualitative questions designed to understand what underlies both positive and negative responses. To our knowledge this is the first mixed-method study about childbirth experiences pre- and during the COVID-19 pandemic. Our research question was: *What were the labour, birth, post-partum and neonatal care experiences of UK and NL women before and during the COVID-19 pandemic and what explains any differences?*

## Methodology

### Study design

This study is based on analysis of the quantitative and qualitative data from the online B3-survey version 3. The B3-survey was designed with help of the EU-funded COST IS1405 network to assess and interpret differences in the quality of maternity care within and between countries. The survey is designed to capture women’s views about what went well in their maternity care experience and what could have worked better.

### Data collection

The B3 survey has been run since February 2014, with an overall response to date of over 97,000 women in 25 languages from 86 countries [15-19]. The third version of the survey has been open from June 2020 (with a planned closure date of June 2022). Women are eligible to complete the survey if they had a baby in the previous three years. For this analysis, responses recorded by women giving birth in the UK and the NL between June and December 2020 were used, encompassing women who gave birth to their most recent baby between 2017 and December 2020. The survey was accessible on the BBB survey website (https://www.babiesbornbetter.org/surveyportal/) and was widely disseminated by researchers, maternity care organisations, service-user organisations, hospitals and midwifery practices through social media from June till September 2020. Responses from women who gave birth in the UK and the NL were collected, but the survey was translated and available in 25 languages, so women could complete the survey in their preferred language, even if this was not English or Dutch.

The survey captures demographics (such as age, parity, self-determined standard of life), clinical factors (such as type of birth and problems during pregnancy). The place name or postcode of the birth location was also captured. Problems during pregnancy were self-reported by the participants and were categorised as *no problems, minor and severe medical and non-medical problems*. Self-reported standard of life was assessed with the following question: ‘*compared to most people in this country, I think my standard of life is…’* followed by a Likert scale comprising the terms *much worse, below average, average, above average and much better*. Women are asked to rate their overall birth experience with the following question: ‘*How do you feel about your labour and birth experience?*’ using a Likert scale of *mostly a very good experience, mostly a good experience, some of it was good and some of it was bad, mostly a bad experience or mostly a very bad experience*.

After that, respondents were asked to record the three most positive aspects of the care they received, and three aspects that could have improved their birth experience. They were then invited to imagine that a close relative or friend who was pregnant had asked for an honest description of the care they had received, and to record what their response would be. The survey ends with an open question where the woman can add any additional information about their labour and birth that they think might be relevant.

### Data analysis

Participants were divided into 2 categories: ‘pre-COVID’ and ‘COVID’. A participant belonged to the ‘pre-COVID’ category if they gave birth before 1 March 2020 and to the ‘COVID’ category if they had a baby from 1 March 2020 onwards. Participants who did not record a date of birth of their baby (n=5) were not included in the analysis.

#### Quantitative analysis

Descriptive analyses were performed on all quantitative data in the survey, firstly by whether the participant gave birth in the UK or the NL, and then by categorising participants into ‘pre-COVID’ and ‘COVID’. Chi-squared tests were performed on categorical variables by COVID status, separately by country and between countries.

The dependent variable ‘birth experiences’ was then dichotomised into two categories: positive and not positive. Positive contained responses ‘mostly good’ and ‘quite good’ and not positive contained responses ‘some good, some bad’, ‘quite bad’, ‘mostly bad’. Standard of life was also dichotomised into ‘below average & average’ and ‘above average’. Birth experiences and standard of life were separately compared in the pre-COVID and COVID time periods and also separately by country, because it is known that a higher than average standard of life might have impacted childbirth experiences during the COVID-19 pandemic [20]. Stata version 17.0 was used for all quantitative data analysis. All statistical tests were carried at the 5% level of significance.

#### Qualitative analysis

We analysed responses to the two open-ended questions in the BBB survey: question 1) “In the place where you gave birth, what were the three most positive experiences of your care?” and question 2) “What do you think could have made your experience better?”. We analysed the answers using Braun and Clarke’s [21] inductive thematic approach. We used this method because it offers flexibility and theoretical freedom to explore experiences, meanings and the reality of participants [21].

First, all the answers were coded with initial codes with the help of MAXQDA version 2020.4.1. These initial codes were used to recognise patterns and were sorted into groups. For each coding group, we then looked at the answers given by women who had given birth before or during the COVID-19 pandemic. The initial themes were developed by LB, and were reviewed and refined through discussions with all authors until consensus was reached. The writing process was guided by the Consolidated Criteria for Reporting Qualitative Research (COREQ) [22]. Since many responses were in Dutch, the quotes were translated by the Dutch-speaking authors collaboratively.

### Reflexivity statement

Our research team comprised seven women with different backgrounds, including midwifery, psychology, statistics and women’s studies. At the beginning of the analysis phase, we all believed that restricting women’s rights during childbirth in an attempt to prevent COVID-19 could lead to short and long-term effects on women’s psychological wellbeing. While prevention of infection is important, lack of attention to psychological wellbeing may generate greater harm than benefit in the longer term [23, 24]. We purposively looked for disconfirming data to ensure that these beliefs did not unduly influence our findings.

### Ethics statement

In the Netherlands the study was submitted to the Medical Ethics Review Committee of the VU University Medical Centre (reference number 2020.255). No ethical approval was needed, since the Medical Research Involving Human Subjects Act did not apply to this study, as there was considered to be no infringement on the physical and/or psychological integrity of the participants.

In the United Kingdom the study was approved by the University of Central Lancashire (UCLan) Committee for Ethics and Integrity (STEMH 449 Amendment_1Jun20).

All the participants gave informed consent before they started the survey. The survey is completely anonymous, so they were aware that they could not withdraw their data once they had submitted their responses.

## Results

### Demographics

A total of 2203 completed surveys from the UK and The NL were recorded between June and December 2020. 1303 (60%) of responses were from the UK and 900 (40%) were from the NL. Overall, 31% (n=678) of respondents gave birth during the COVID-19 pandemic (March-December 2020); 36% (468) of all UK respondents, and 23% (n=210) of all respondents from the NL [Table 1].

**Table 1.** Babies Born Better Survey responses in relation to COVID-19 pandemic

|  | UK | Netherlands | Total |
| --- | --- | --- | --- |
| **Responses** | 1303 | 900 | 2203 |
| **Baby during COVID-19; n (%)\*\*** |  |  |  |
| Yes | 468 (36.0) | 210 (23.4) | 678 (30.9) |
| No | 833 (64.0) | 687 (76.6) | 1520 (69.1) |
| **Year of birth: n (%)\*\*** |  |  |  |
| 2017 | 89 (7.0) | 86 (9.8) | 175 (8.1) |
| 2018 | 301 (23.6) | 218 (24.7) | 519 (24.1) |
| 2019 | 349 (27.4) | 311 (35.3) | 660 (30.6) |
| 2020 | 535 (42.0) | 267 (30.3) | 802 (37.2) |

+March 2020 to Dec 2020
\* 5 missing responses
\*\* 47 missing responses

Table two provides demographic details [Table 2]. Median age (IQR) was 31 years old in the UK and 32 years old in the NL with minimal difference in the COVID periods. Highest level of education also showed minimal differences between COVID periods for both countries. However, the NL had a higher percentage (21%) of participants completing tertiary education compared to the UK (11%). No significant differences were found in self-reported standard of life in either UK or the NL pre- or during the pandemic.

**Table 2.** Demographics of participants who completed the survey

|  | **UK** |  |  | **Netherlands** |  |  |
| --- | --- | --- | --- | --- | --- | --- |
|  | **Pre-COVID** | **COVID** | **All** | **Pre-COVID** | **COVID** | **All** |
| **Age (years old); median (LQ, UQ)** | 31 (28,34) | 31 (28,34) | 31 (28,34) | 32 (29,34) | 31 (28,34) | 32 (29, 34) |
| **Educational level; n (%)** | **Pre-COVID** | **COVID** | **All** | **Pre-COVID** | **COVID** | **All** |
| No formal schooling | 1 (0.1) | 1 (0.2) | 2 (0.2) | 1 (0.2) | 0 | 1 (0.1) |
| Primary education | 1 (0.1) | 1 (0.2) | 2 (0.2) | 1 (0.2) | 1 (0.5) | 2 (0.2) |
| Secondary education | 77 (9.3) | 68 (14.3) | 145 (11.2) | 11 (1.6) | 1 (0.5) | 12 (1.3) |
| Tertiary/professional/technical | 86 (10.4) | 50 (10.5) | 136 (10.5) | 129 (19.1) | 57 (25.6) | 186 (20.7) |
| University or equivalent | 659 (80.0) | 357 (74.8) | 1016 (78.1) | 535 (79.0) | 164 (73.5) | 699 (77.7) |
| **p-value** |  |  | **p=0.097** |  |  | **p=0.145** |
| **Standard of life; n (%)** | **Pre-COVID** | **COVID** | **All** | **Pre-COVID** | **COVID** | **All** |
| Much worse | 0 | 0 | 0 | 1 (0.2) | 0 | 1 (0.1) |
| Below average | 15 (1.8) | 3 (0.6) | 18 (1.4) | 13 (1.9) | 2 (0.9) | 15 (1.7) |
| Average | 352 (42.7) | 201 (42.1) | 553 (42.5) | 261 (38.6) | 99 (44.4) | 360 (40.0) |
| Above average | 385 (46.7) | 226 (47.3) | 611 (46.9) | 363 (53.7) | 108 (48.4) | 471 (52.4) |
| Much better | 72 (8.7) | 48 (10.0) | 120 (9.2) | 38 (5.62) | 14 (6.3) | 52 (5.8) |
| **p-value** |  |  | **p=0.293** |  |  | **p=0.433** |

In the UK, 46% (n=380) reported pregnancy problems pre-COVID and 40% (n=193) during COVID. There was a weak statistically significant difference in the UK (X2 (1) = 4.2, p = .041) but no difference was found in the NL [Table 3].

**Table 3.** Problems in pregnancy of the included participants

|  | **UK** |  |  | **Netherlands** |  |  |
| --- | --- | --- | --- | --- | --- | --- |
|  | **Pre-COVID** | **COVID** | **All** | **Pre-COVID** | **COVID** | **All** |
| **Problems in pregnancy; n (%)** | 380 (46.1) | 193 (40.3) | 573 (44.0) | 273 (40.3) | 88 (39.5) | 361 (40.1) |
| **p-value** |  |  | **p=0.041\*** |  |  | **p=0.820** |
| **Type of problems; n (%)** | **Pre-COVID** | **COVID** | **All** | **Pre-COVID** | **COVID** | **All** |
| Minor non-medical problems | 82 (21.2) | 36 (18.1) | 118 (20.1) | 69 (24.6) | 20 (22.2) | 89 (24.0) |
| Minor medical problems | 243 (62.8) | 138 (69.4) | 381 (65.0) | 167 (59.4) | 57 (63.3) | 224 (60.4) |
| Severe non-medical problems | 15 (3.9) | 3 (1.5) | 18 (3.1) | 5 (1.8) | 2 (2.2) | 7 (1.9) |
| Severe medical problems | 47 (12.1) | 22 (11.1) | 69 (11.8) | 40 (14.2) | 11 (12.2) | 51 (13.8) |
| **p-value** |  |  | **p=0.258** |  |  | **p=0.897** |

\* $p \leq 0.05$

In the NL women reported a higher percentage of having a vaginal birth without help in both time periods compared to the UK. No statistically significant difference was found in either country between COVID time points for type of birth [Table 4]. There were statistically significant differences (*X*2 (4) = 15.38, *p* = .004) in birth setting in the UK with more women giving birth in hospital during COVID-19 pandemic compared to pre-COVID. No such difference was found in the NL, but the NL had a higher rate of homebirth overall among the respondents: 44% (n=399) compared to the UK; 18% (n=230).

**Table 4.**
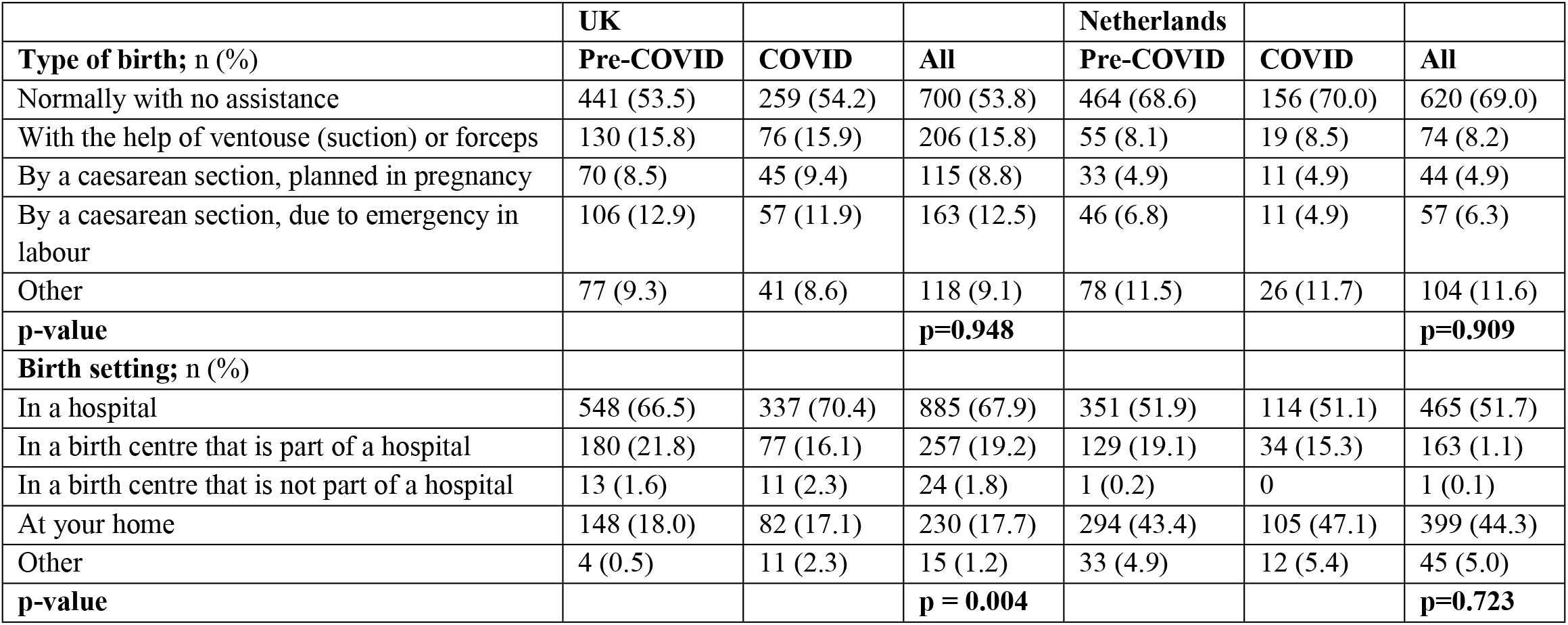
Type of birth and birth setting of the included participants

Most of the described demographics are in line with the population numbers, but the home birth rate in our sample is higher than in the overall population, for both the UK and the NL [25, 26].

### Quantitative data

The NL had a higher percentage of women reporting a very good birth experience 65% (n=581) compared to the UK 48% (n=624), which is a statistically significant difference between the two countries (p-value < 0.01). No significant differences were found in either country for labour and birth experiences pre and during the COVID-19 pandemic for the respondents as a whole [Table 5]. At both timepoints, and for those reporting all standards of life, reports of a positive experience were statistically significantly higher in the NL than in the UK.

**Table 5.** Overall birth experience of the participants

|  | UK |  |  | Netherlands |  |  |
| --- | --- | --- | --- | --- | --- | --- |
| Labour & Birth experience; n (%) | Pre-COVID | COVID | All | Pre-COVID | COVID | All |
| It was mostly a very good experience | 380 (46.2) | 244 (51.2) | 624 (48.0) | 425 (62.9) | 156 (70.0) | 581 (64.6) |
| It was mostly quite a good experience | 149 (18.1) | 82 (17.2) | 231 (17.7) | 138 (20.4) | 43 (19.3) | 181 (20.1) |
| Some of it was good, some of it was bad | 164 (19.9) | 100 (21.0) | 264 (20.3) | 69 (10.2) | 16 (7.2) | 85 (9.5) |
| It was mostly quite a bad experience | 74 (9.0) | 31 (6.5) | 195 (8.1) | 26 (3.9) | 5 (2.2) | 31 (3.4) |
| It was mostly a very bad experience | 56 (6.8) | 20 (4.2) | 76 (5.9) | 18 (2.7) | 3 (1.4) | 21 (2.3) |
| p-value within country | p=0.103 |  |  | p=0.237 |  |  |
| p-value between countries overall | P < 0.01 |  |  |  |  |  |

In the UK, similar rates of women reporting average and below standard of life had positive labour and birth experiences pre-COVID 64% (n=233) compared to during the COVID-19 pandemic (65%, n=132). Women reporting a higher standard of life were non-significantly more likely to report better experiences if they gave birth during the pandemic than those who gave birth before COVID (71% vs 65%).

In the NL, similar patterns were seen, with the more positive birth experience for women who gave birth during the pandemic reaching statistical significance for women self-reported higher standard of life (94% vs 84%) [Table 6].

**Table 6.**
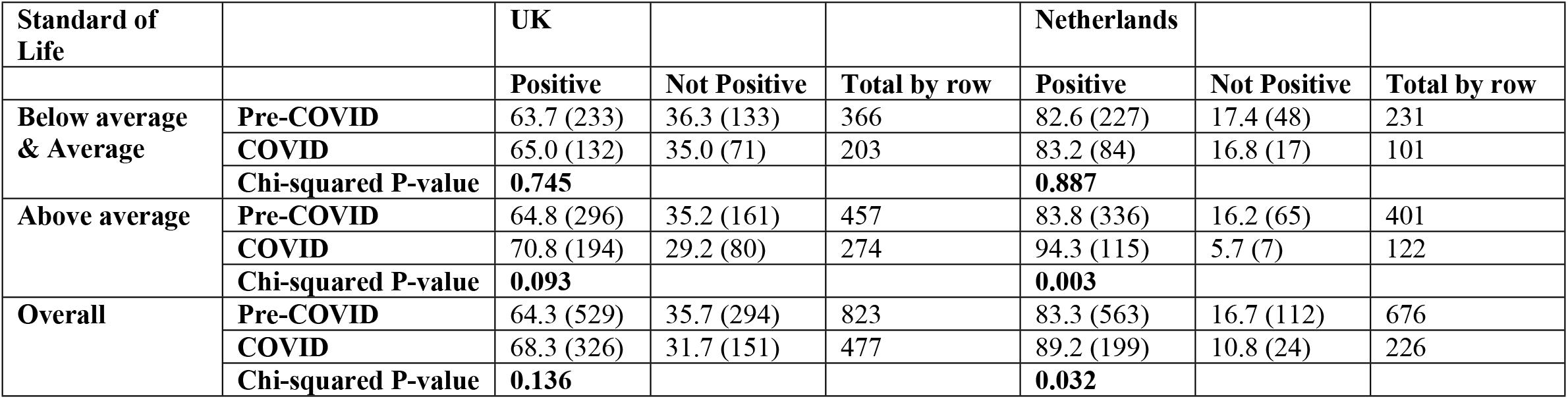
Standard of Life and childbirth experiences of the included participants

| **Standard of Life** |  | **UK** |  |  | **Netherlands** |  |  |
| --- | --- | --- | --- | --- | --- | --- | --- |
|  |  | **Positive** | **Not Positive** | **Total by row** | **Positive** | **Not Positive** | **Total by row** |
| **Below average & Average** | **Pre-COVID** | 63.7 (233) | 36.3 (133) | 366 | 82.6 (227) | 17.4 (48) | 231 |
|  | **COVID** | 65.0 (132) | 35.0 (71) | 203 | 83.2 (84) | 16.8 (17) | 101 |
|  | **Chi-squared P-value** | **0.745** |  |  | **0.887** |  |  |
| **Above average** | **Pre-COVID** | 64.8 (296) | 35.2 (161) | 457 | 83.8 (336) | 16.2 (65) | 401 |
|  | **COVID** | 70.8 (194) | 29.2 (80) | 274 | 94.3 (115) | 5.7 (7) | 122 |
|  | **Chi-squared P-value** | **0.093** |  |  | **0.003** |  |  |
| **Overall** | **Pre-COVID** | 64.3 (529) | 35.7 (294) | 823 | 83.3 (563) | 16.7 (112) | 676 |
|  | **COVID** | 68.3 (326) | 31.7 (151) | 477 | 89.2 (199) | 10.8 (24) | 226 |
|  | **Chi-squared P-value** | **0.136** |  |  | **0.032** |  |  |

### Qualitative data

Given that the findings that the experiences of women were similar or better during Covid than pre-Covid are in direct contrast to many other studies in this area, we examined the responses to two open questions to try to identify reasons for our unexpected results. Our thematic analysis identified two themes underpinning negative experiences: 1) Lack of support; 2) Limits placed on freedom of choice. Two further themes might explain why some reported experiences were more positive than anticipated: 3) Lower expectations of care during the pandemic; and 4) Efforts of staff to give personalised care, despite the rules. All four themes were more present in the accounts of women who gave birth during the pandemic.

### Lack of support

Responses to the question ‘What do you think could have made your experience better?’ indicate that women who gave birth during the COVID-19 pandemic revealed a lack of support before, during and after childbirth. For some women this was associated with fewer staff being available, or by shorter or more distant contacts with staff due to COVID-19 social distancing measures

> *‘When I unexpectedly laboured I did not receive enough care waiting for my COVID test and labouring waiting for C section. ’ (‘What do you think could have made your experience better?’, UK woman, second child, gave birth August 2020)*
>
> *‘More presence of the midwife and nurse during childbirth’ (‘What do you think could have made your experience better?’, NL woman, first child, gave birth July 2020)*

Furthermore, COVID-19 measures made it more difficult for women to receive their chosen labour and post-partum support from a partner, family member and/or doula, due to restrictions in both countries.

> *‘Because of the corona measures my partner was not allowed to be there during overnight stay before the birth’ (‘What do you think could have made your experience better?’, NL woman, first child, gave birth April 2020)*
>
> *‘Partner being able to stay longer on the ward - limited visiting due to covid’ (‘What do you think could have made your experience better?’, UK woman, second child, gave birth September 2020)*
>
> *‘Other family being able to visit the post-natal ward’ (‘What do you think could have made your experience better?’, UK woman, second child, gave birth September 2020)*

### Limits placed on freedom of choice

The majority of the participants considered freedom to make choices about their labour and birth to be very important. While many women reported that their freedom to make choices was respected during childbirth, even during the COVID-19 pandemic, some of those giving birth during restrictions noted limitations to this freedom. This was particularly noted in terms of choices about place of labour and birth in the UK, due to some home birth services being closed.

> *‘Home birth service’ (‘What do you think could have made your experience better?’, UK woman, first child, gave birth June 2020)*

None of the respondents from the NL noted restrictions in home birth services, and for some, freedom to give birth at home during the pandemic was a positive factor in their experience:

> *‘That I could give birth at home’ (‘In the place where you gave birth, what were the three most positive experiences of your care?’, NL woman, third child, gave birth May 2020)*

Furthermore, some respondents experienced limitations in other parts of their birth experience, including the possibility to choose a water birth:

> *‘Due to Covid I was not allowed a water birth. ’ (‘What do you think could have made your experience better?’, UK woman, first child, gave birth April 2020)*

### Lower expectations of care during the pandemic

As the previous results have shown, many of the women who gave birth during the COVID-19 pandemic in both the UK and the NL were alone when they wanted companionship (family and/or professional), and had less freedom of choice. This being the case, it is surprising that the quantitative data suggest that they were equally, or even more likely to report that their birth was a positive experience compared to women who gave birth before the pandemic. One of the reasons behind this might be that, during the pandemic, women had lower expectations for their labour and birth experience, due to national and local restrictions. For example, the following examples were cited as distinct positives of care by two women giving birth during the pandemic, even though this would be seen as basic care standard in pre-pandemic times

> *‘My own [primary care] midwife could attend the birth’ (‘In the place where you gave birth, what were the three most positive experiences of your care?’, NL woman, first child, gave birth August 2020)*
>
> *‘Flexibility so I could be supported by my husband in a stressful situation’ (‘In the place where you gave birth, what were the three most positive experiences of your care?’, UK woman, first child, gave birth August 2020)*

### Efforts of staff to give individualised care, despite the rules

Many women in both groups, pre-COVID and COVID, valued the importance of known, competent, caring and empathic care providers. The women who gave birth during the COVID-19 pandemic tended to be complimentary about their care providers, particularly in relation to their efforts at a difficult time.

> *‘The doctors and nurses were very helpful and flexible (despite Corona measures)’ (‘In the place where you gave birth, what were the three most positive experiences of your care?’, NL woman, first child, gave birth March 2020)*

Several women were grateful for maternity care providers making exceptions to the COVID measures.

> *‘This was particular to the time of our birth, when there were still many Covid-19 related restrictions in place, but the healthcare providers allowed my partner to be there from the beginning of my induction drip rather than waiting until 4cm dilated which I know was the case in other Trusts/hospitals’ (‘In the place where you gave birth, what were the three most positive experiences of your care?’, UK woman, first child, gave birth May 2020)*

One participant acknowledged that her childbirth experience during the pandemic was challenging, but complimented her midwives that they made the experience slightly better.

> *‘My midwives were very supportive and helpful. They [my midwives] made giving birth during a pandemic slightly better’ (‘In the place where you gave birth, what were the three most positive experiences of your care?’, UK woman, first child, gave birth April 2020)*

## Discussion

In this multi-method study we compared the childbirth experiences of UK and NL women responding to the international B3 survey who gave birth before and during the COVID-19 pandemic. In general, women giving birth in the NL were much more likely to rate their care as positive than women giving birth in the UK, at both time points, and across all self-report standard of life categories. In each country, women who gave birth during the COVID-19 pandemic were, on average, at least as positive about their childbirth experiences as women who gave birth before the COVID-19 pandemic.

Moreover, the NL women who self-reported an above average standard of life were even more positive about their birth experiences if they gave birth during the COVID-19 pandemic, and while this trend was also seen in the UK data, it did not reach statistical significance. This is despite the fact that many women labouring during the pandemic reported having less support, choice and control than those in the pre-pandemic period. Based on the qualitative analyses, two mitigating factors appeared to contribute to this unexpected finding: women seemed to have lower expectations during the pandemic, and they appeared to appreciate that care providers tried hard to personalise care, sometimes despite the rules they were supposed to be following.

### Interpretation and comparison with other literature

This study contributes to the ongoing evidence that the COVID-19 pandemic has had an impact on women’s labour, birth, post-partum and neonatal care experiences, and the mental health of women, partners, and their babies [27-29]. Women in our study reported that they could not get the support they wanted, either from their maternity care providers or from their chosen birth partner(s), because of COVID-19 measures. Other researchers have linked the lack of family and healthcare professional support to feelings of loneliness, especially where it continues into the post-partum period [27, 30]. The importance of choice in matters such as place of labour and birth, water birth, and birth partners for women is well documented [31-34]. Many of our respondents wanted options to choose for homebirth and community services, before and during the pandemic. A study in the United States revealed that women’s increased interest for community care during the pandemic may have been due to hospital being perceived as a less safe option [35]. Differences between the UK and NL in access to home birth services is likely due to homebirth services being maintained in the NL during the pandemic. In contrast in the UK, several home birth and midwife led unit services were closed and maternity care was centralised into hospitals to concentrate staffing in the acute units [14].

Despite the restrictions to companionship and freedom to choose, our quantitative data indicate that survey respondents who laboured and gave birth during the pandemic phase were, on average, at least as positive about their labour and birth experiences as those giving birth before this point. This is in direct contrast to most surveys to date [36, 37]. The responses to the open questions in the survey shed some light on this phenomenon. Many women reported they were particularly grateful for staff who went the extra mile to ensure personalised care, sometimes by making exceptions, even though they were often not praised for it [38]. While this was appreciated and beneficial for many respondents, there is growing evidence of moral distress and compassion fatigue among staff who are trying to maintain services by continuously having to go ‘above and beyond’ their shift times, or by the stress of breaking rules that they feel are damaging to women, birthing people and families [39, 40]. This is not sustainable and could be highly detrimental to staff wellbeing and therefore to safe and personalised care in the longer term [41]. Moreover, there is an equity problem if women in higher socio-economic groups are more likely to benefit from this. Our study highlights the inequities during crisis time. The NL participants with a self-reported higher than average life standard had higher birth satisfaction during the COVID-19 pandemic than women with a self-reported average or lower than average life standard. These findings are confirmed by the study by Mollard and Kupzyk [20]. A hypothesis for this difference might be that women and their families with an above average standard of life had, in general, less stressors during the pandemic, such as job insecurity, loss of income and caregiving burden [42]. It could also be that, in a crisis situation where resources and emotional capacity are limited, staff respond even more to those who are most able to articulate their need for individualised care compared to non-crisis situations [43]. Personalisation should continue to be an important part of general policy in maternity care, and therefore of care provision at all levels of the service, including guidelines and staffing resources, to enable staff to benefit all service users equitably.

### Strengths and limitations of this study

The strengths of our study include the large number of responses to the B3 survey both before and after the pandemic, the ability to compare birth experiences between two European countries with similar social structures, and the use of both quantitative and qualitative data. The combination of methods gave us the opportunity to explain some of the unexpected findings in the quantitative data. As far as we are aware there are no other published studies to date that can make this comparison for women using childbirth services before and after the pandemic. Furthermore, our team consisted of people from a range of different disciplines, which contributed to investigator triangulation and therefore the trustworthiness of the results.

However, our study has some limitations. Since the data were downloaded from the survey platform in December 2020, more women who gave birth during COVID-19 responded soon after their birth than women who gave birth pre-pandemic. This might have led to different levels of recall bias between the two groups. Although memory for the actual events of labour and birth is generally stable over time, there is some evidence that affective memories may become less positive, particularly if the labour or birth were difficult [44]. Furthermore, we used a convenience sampling method since we disseminated the survey online. Our results are therefore relevant to women who have similar characteristics to those in our sample. As our data suggest, women who self-identify as having a lower than average standard of life are more likely to report negative labour and birth experiences. Most studies report a disproportionately high adverse impact of the pandemic on women who were from a minority background, or those with low health literacy [24]. These women may also have been missing from our sample. Even though the B3 survey is available in 25 languages, this would not benefit women who do not read their written language, or who do not have access to the survey on smart phones or computers. Women having home births were overrepresented in both countries. However, this does not explain the higher percentage of positive responses in the NL, since women having home births tend to be more positive in their responses, so the over-representation of home births in the UK suggests that, with a representative sample from both countries, these differences would have been even greater.

## Conclusion

In this study, respondents in both the UK and the NL who gave birth during the pandemic were, on average, as likely, or more likely to rate their labour and birth experience positively when compared to women who gave birth pre-pandemic. This was despite the fact respondents who gave birth during the pandemic reported lack of support and choice. This apparent paradox may be partly because women had fewer expectations of the care they were going to receive, which made them value normal care practices more. There is also evidence that they valued the work of maternity care providers who persisted with giving individualised care during the COVID-19 pandemic, despite the constraints of resources and rules, and that this influenced positive care ratings. Our study implies that women were adversely affected by COVID-19 restrictions during their labour and birth, but that there were mitigating factors for some women, especially those with an above average self-reported standard of life. Equitable personalisation of care is important during a crisis as well as in normal times, and should be part of general policy in maternity care and routine implementation in actual practice to benefit all service users, rather than being maintained for some, and, even then only through the willingness of staff to go ‘above and beyond’.

## Data Availability

The Babies Born Better survey data will be available from the Data Services (ReShare data repository) and DOIs will be provided on paper acceptance.

## Acknowledgements

We are grateful to all the participants for their willingness to share their experiences with us. We acknowledge the work of those who contributed to developing the Babies Born Better Survey. Details of the project, the Steering Group, and the Country Coordinators can be found at http://www.babiesbornbetter.org/about/. Furthermore, we would like to thank maternity care organisations in the NL and the UK for sharing the Babies Born Better Survey, and the ASPIRE COVID-19 research team for their input during the study.

## Author contributions

### Conceptualisation

Lauri M.M. van den Berg, Naseerah Akooji, Gill Thomson, Ank de Jonge, Marie-Clare Balaam, Anastasia Topalidou and Soo Downe.

### Data curation

Lauri M.M. van den Berg and Naseerah Akooji.

### Formal analysis

Lauri M.M. van den Berg, Naseerah Akooji, Gill Thomson, Ank de Jonge, Marie-Clare Balaam, Anastasia Topalidou and Soo Downe.

### Investigation

Naseerah Akooji.

### Methodology

Lauri M.M. van den Berg, Naseerah Akooji, Gill Thomson, Ank de Jonge, Marie-Clare Balaam, Anastasia Topalidou and Soo Downe.

### Project administration

Soo Downe.

### Resources

Gill Thomson, Ank de Jonge, Anastasia Topalidou and Soo Downe.

### Supervision

Gill Thomson, Ank de Jonge and Soo Downe.

### Visualisation

Lauri M.M. van den Berg and Naseerah Akooji.

### Writing – original draft

Lauri M.M. van den Berg and Naseerah Akooji.

### Writing – review & editing

Lauri M.M. van den Berg, Naseerah Akooji, Gill Thomson, Ank de Jonge, Marie-Clare Balaam, Anastasia Topalidou and Soo Downe.

All authors read and approved the final manuscript.

## Funding

This research is funded by the Economic and Social Research Council (ESRC), as part of UK Research and Innovation’s rapid response to COVID-19 [grant number ES/V004581/1]. Full details of the main study are available via ResearchRegistry (researchregistry5911) and via UKRI Gateway (https://gtr.ukri.org/projects?ref=ES%2FV004581%2F1).

This research article was based on the Babies Born Better project that was developed as part of the EU-funded COST Action IS1405: BIRTH: ‘Building Intrapartum Research Through Health - an interdisciplinary whole system approach to understanding and contextualising physiological labour and birth,’ sustained by the COST (European Cooperation in Science and Technology) Programme as part of EU HORIZON 2020.

## Conflict of interest

The authors declare that they have no competing interests.

## Notes

### Competing Interest Statement

The authors have declared no competing interest.

### Author Declarations

In the Netherlands the study was submitted to the Medical Ethics Review Committee of the VU University Medical Centre (reference number 2020.255). No ethical approval was needed, since the Medical Research Involving Human Subjects Act did not apply to this study, as there was considered to be no infringement on the physical and/or psychological integrity of the participants. In the United Kingdom the study was approved by the University of Central Lancashire (UCLan) Committee for Ethics and Integrity (STEMH 449 Amendment_1Jun20). All the participants gave informed consent before they started the survey. The survey is completely anonymous, so they were aware that they could not withdraw their data once they had submitted their responses.

